# A dose scaling antivenin protocol in treatment of *Daboia palaestinae* envenomation may reduce morbidity and costs

**DOI:** 10.1101/2025.01.29.25321324

**Authors:** Daniel J. Jakobson, Zurab Zakariashvili, Enzo F. Galicia H., Mohammad Abu Issa, Miguel M. Glatstein, Frederic S. Zimmerman

## Abstract

**Background:** *Daboia palaestinae* is a leading cause of snakebite envenomation in the eastern Mediterranean, with substantial mortality in absence of antivenin. Current recommended antivenin dose is 50 ml; however, antivenin is costly, may be difficult to obtain and is associated with substantial morbidity. Thus, this study was designed to define the minimal effective antivenin dose and identify patients who can be safely treated without antivenin.

**Methods:** This retrospective single-center study was conducted in adults with suspected or confirmed D. *palaestinae* envenomation. Patients were treated via our previously developed envenomation protocol: no antivenin use for local symptoms and dose scaling for mild or severe systemic symptoms – initially 10 ml antivenin, with repeat dosing for ongoing systemic symptoms. Main outcomes measured were morbidity and mortality associated with this protocol. Secondary outcomes included assessing the demographics and clinical effects of snake envenomation and comparing between those who received antivenin and those who did not.

**Results:** 101 patients were included. 45 minutes [median; interquartile range: 30-61 minutes] elapsed between envenomation and hospital admission, with no differences between groups. Among 52 patients receiving antivenin, 119 [60-237] minutes elapsed between envenomation and initial antivenin administration, with a maximum of 1073 minutes to initial anvenin administration. Maximum until last antivenin was 3860 minutes.

Median antivenin dose was 15 [10-22.5] ml, with 26/52 (50.0%) requiring only 10 ml. 2 patients developed an early antivenin immune reaction, with 1 developing anaphylaxis requiring invasive ventilation. No patients died during hospitalization.

**Conclusions:** This cohort demonstrates that a dose-scaling antivenin protocol can be safely employed, reducing morbidity and costs. This suggests a randomized control trial comparing fixed dose regimen to an escalation protocol and development of similar protocols for envenomations due to other snake species.

## Introduction

Snake envenomation is a widespread cause of mortality and morbidity worldwide, with snake bites resulting in 50 – 130,000 deaths annually. Native species of venomous snakes are found on all continents except Antarctica, whereas non-native snakes may also be encountered as pets [1, 2]. Clinical signs and symptoms, as well as symptom severity, defer between and within species and are dependent on the chemical composition of the venom, which varies substantially between species, with intraspecies variation occurring as well [2, 3].

*Daboia palaestinae (*previously *Vipera palaestinae* or *Vipera xanthina palaestinae*) is a viper species endemic to the eastern coasts of the Mediterranean, especially the coastal plains and inland hills of Israel and Lebanon, and is the most common poisonous snake and the leading causing of snakebite envenomation in this region [3, 4], with 100-300 snakebites reported in Israel annually [5, 6]. Taxonomically, it is from the Viperinae subfamily, which is distributed widely across Africa, Europe, the Middle East and Asia, including the islands of the far east [3].

The venom of this snake primarily contains myotoxins, procoagulants and hemorrhagins, as well as neurotoxins, angioneurin growth factors, and integrin inhibitors which can cause both local and systemic effects [3-5, 9]. Local effects include severe pain and swelling of the affected limb, whereas systemic affects include nausea, vomiting and abdominal pain, with more severe envenomizations resulting in respiratory distress, hypotension and convulsions, up to hypovolemic and vasodilatory shock which, left untreated, can lead to cardiac arrest. Neurological symptoms, coagulopathy, thrombocytopenia and necrosis have also been reported [4, 8-10]. Effects may appear immediately after snake bite, but are often delayed, including beyond 24 hours [9].

The primary and most effective treatment of snake bite envenomation, including that resulting from *D. palaestinae* bite, is administration of specific or polyvalent snake antivenin, use of which drastically reduces mortality, with a reduction in morbidity as well [8, 9].

Currently, a fixed dose regimen of 50 ml of antivenin is recommended in all *D. palaestinae* envenomations, including both systemic manifestations and progressive local manifestations of envenomation where *D. palaestinae* bite is suspected or confirmed. However, antivenin administration is costly, with a single fixed dose regimen estimated cost of approximately $7000, with repeat administration sometimes required [4, 5, 11]. Furthermore, certain antivenins have historically been in short supply [11]. Moreover, antivenin administration can itself be associated with substantial morbidity, including anaphylaxis, chills, fever, pain at injection site and serum sickness, including delayed reactions up to a month after administration [4, 8, 10].

Thus, though antivenin administration is absolutely essential in preventing morbidity and mortality in the relevant patient population, it is also important to define the minimal necessary dose to effectively treat envenomation, as well as define patient populations who can be safely treated without antivenin administration. The achievement of such objectives will minimize antivenin side effects, reduce costs and help to increase the availability of antivenin for those who truly need it.

In contrast to this fixed-dose regimen, we previously developed an envenomation treatment protocol that minimized use of antivenin in order to minimize side effects and overall morbidity, as well as reduce antivenin use in periods of scarcity [10]. This protocol involves conservative treatment of local symptoms without use of antivenin [5, 6, 8] and treatment of both mild and severe systemic symptoms employing dose scaling antivenin, with an initial 10 ml dose of antivenin, with repeat dosing in patients with ongoing systemic symptoms, in accordance with treatment response. This protocol has the potential to reduce antivenin morbidity as well as substantially reduce costs and increase antivenin availability [10]. It should be noted that since initial publication of this escalation protocol, quality control of antivenin production has improved, possibly contributing to reduced morbidity from antivenin administration [1, 4, 11]. Furthermore, we have subsequently further reduced initial antivenin dosing relative to the initial publication.

Thus, the primary aim of the current study is to describe the outcomes of a snake envenomation treatment using an escalated dose antivenin protocol in terms of length of stay, morbidity and mortality. Secondary aims include assessing the demographics and clinical effects of snake envenomation and comparing between those who received antivenin and those who did not.

## Materials and Methods

This retrospective single-center study was conducted at Barzilai University Medical Center, a 500-bed hospital in Ashkelon, Israel. Patients were initially admitted to the intensive care unit, a 12-bed mixed ICU with both medical and surgical patients. The study was approved by the Barzilai University Medical Center institutional review board prior to initiation (approval number: 0009-23-BRZ). Due to the retrospective nature of the study, informed consent was waived.

Study subjects included all adult patients hospitalized in Barzilai University Medical Center from 2014-2023 with an admission diagnosis of snake bite (ICD-10 code T63.0) in which D. *palaestinae* envenomation was suspected or confirmed. Patients admitted with a diagnosis of snake bite in which D. *palaestinae* envenomation was ruled out were excluded.

Admitted patients were preferentially treated according to the protocol currently being applied in our medical center. Thus, patients exhibiting systemic signs of envenomation – including, but not limited to, nausea, vomiting, abdominal pain, diarrhea, blood pressure changes, arrhythmias, and anaphylactic shock – were immediately administered 10 ml of antivenin. Further doses of 10 ml of antivenin were administered if abatement of symptoms was not noted within a short period of time or if symptoms recurred. Patients with local symptoms and signs only did not receive antivenin. All patients were kept under observation in the intensive care unit.

Data were accessed for research until September 12, 2023 and included date and time of envenomation, age, sex, time of presentation to the hospital, clinical signs and symptoms as well as laboratory results on presentation and during hospitalization, time and dosage of antivenin administration, need for repeat administration, complications and complication management, as well as need for adjunctive therapy. Length of stay in the intensive care unit and in-hospital was also recorded. Symptoms and signs were classified as local (confined to the limb affected, e.g. swelling, pain, discoloration), mild systemic (e.g. gastrointestinal symptoms) and severe systemic (e.g. neurological or cardiovascular compromise). All subject identifiers were removed by subject coding prior to data analysis, thus authors had access to data individual participant information during data collection but not after collection and during analysis.

The primary outcome of this study was in-hospital morbidity and mortality associated with an escalated dose antivenin protocol as measured by clinical signs and symptoms as well as laboratory results. Secondary outcomes included evaluation and description of the demographics and clinical effects of snake envenomation, including comparisons between those who received antivenin and those who did not.

Descriptive statistics (i.e. numbers, proportion and means) were used to describe the study population. A secondary analysis compared subjects with local symptoms and signs only – not treated with antivenin by our protocol – to those with systemic symptoms or signs.

Categorical variables were compared using either the Chi square test or the a-parametric Fisher’s exact test as appropriate. Continuous variables were compared using either the Students T-test or the a-parametric Mann-Whitney U tests as appropriate.

The data was downloaded to a Microsoft Excel (Ver. 2019) database and then transferred to R (Version 4.2.2. R Foundation for Statistical Computing. released 2022. Vienna, Austria), which was used for analysis. Comparison between proportions was performed using the χ^2^-score or the Fisher’s exact test. To compare continuous variables the Student’s t-test or the Mann-Whitney-Wilcoxon test was used. In all tests, two-tailed p-values were taken and a p-value <0.05 was considered significant.

## Results

During the study period, 114 patients were admitted to Barzilai University Medical Center with a snake bite diagnosis. After chart review, 10 patients were eliminated after ruling out *D. palaestinae* envenomation, 2 more patients were eliminated due to initial treatment in another hospital and 1 minor patient was eliminated. Thus, 101 patients were included in the current study. Of these, 49 patients did not receive antivenin and 52 received antivenin (Fig 1).

**Fig 1.**
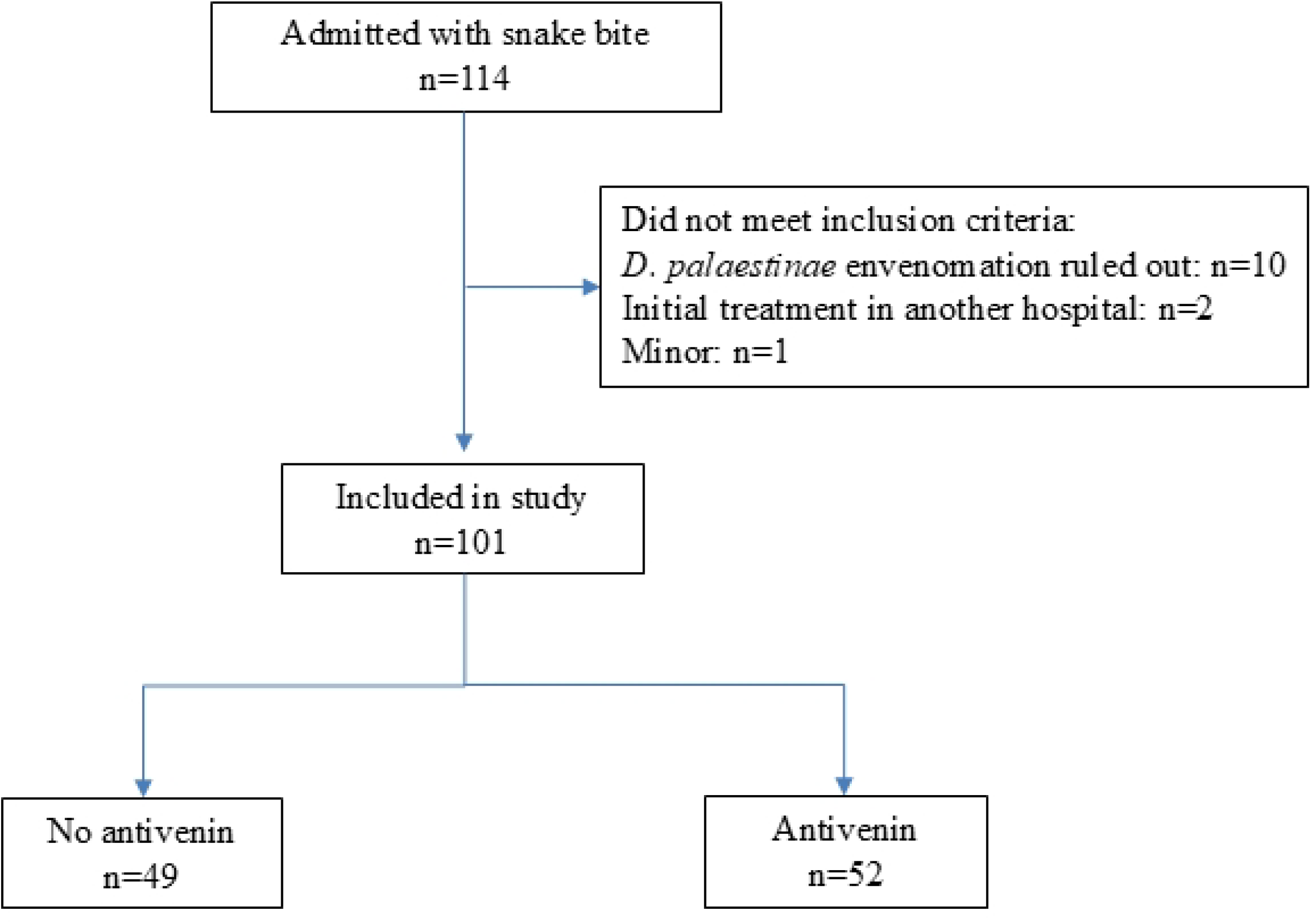
Inclusion / exclusion criteria

The average age of the study population was 41±5 years. 78/101 (77.2%) of patients were male. 24/101 (23.8%) had had a chronic condition. No demographic differences were noted between those who did and did not receive antivenin (Table 1).

**Table 1.**
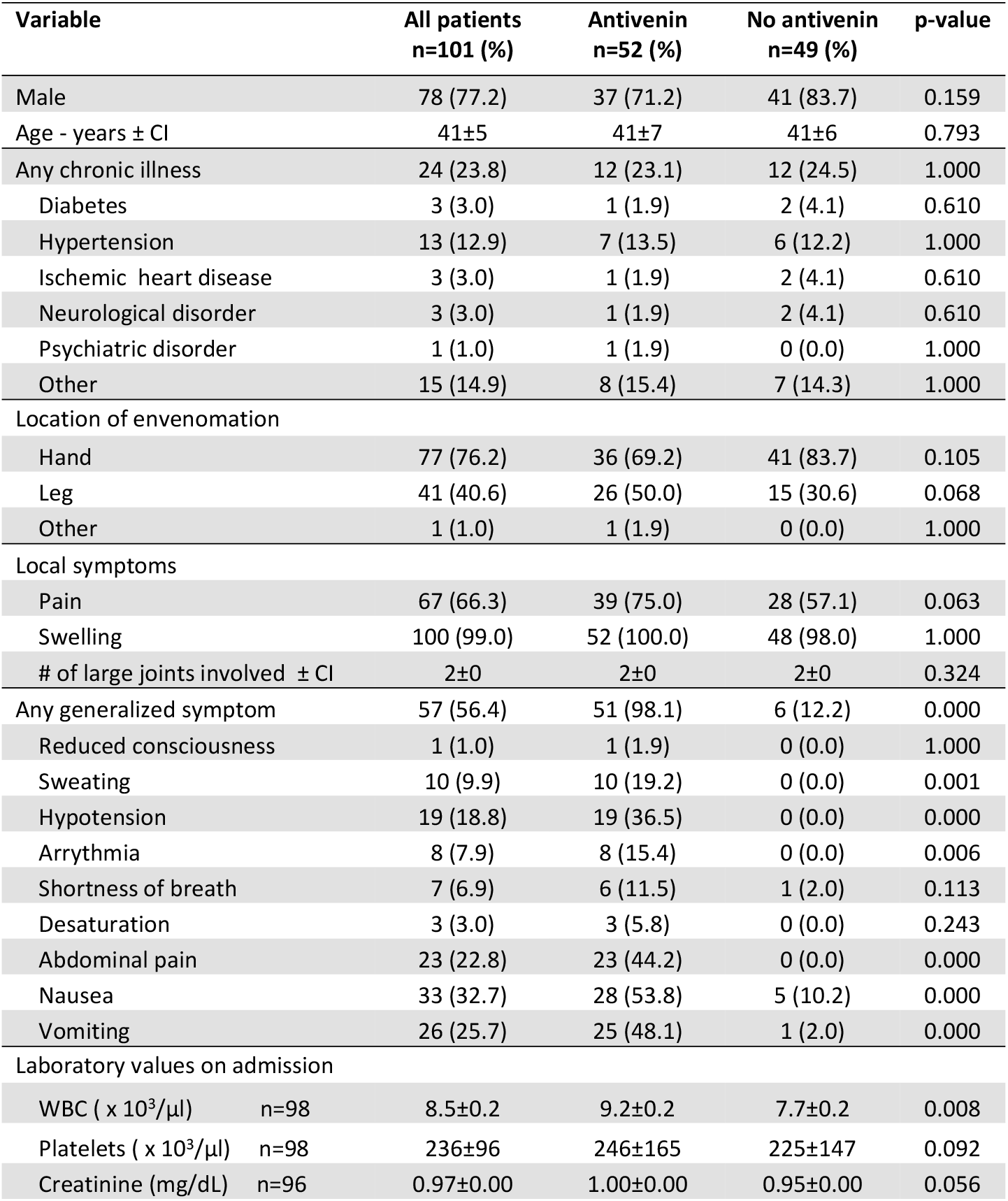
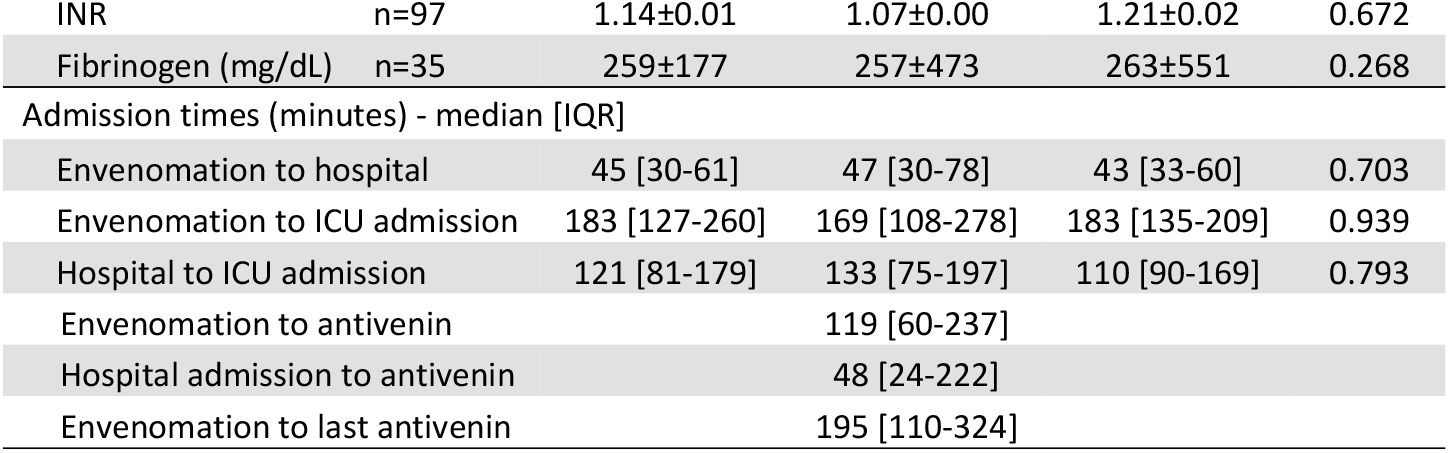
Demographics and presentation. CI: confidence interval; ICU: intensive care unit; <u>INR: international normalized ratio; IQR: interquartile range; WBC: white blood cells.</u>

| Variable | All patients<br>n=101 (%) | Antivenin<br>n=52 (%) | No antivenin<br>n=49 (%) | p-value |
| --- | --- | --- | --- | --- |
| Male | 78 (77.2) | 37 (71.2) | 41 (83.7) | 0.159 |
| Age - years ± CI | $41 \pm 5$ | $41 \pm 7$ | $41 \pm 6$ | 0.793 |
| Any chronic illness | 24 (23.8) | 12 (23.1) | 12 (24.5) | 1.000 |
| Diabetes | 3 (3.0) | 1 (1.9) | 2 (4.1) | 0.610 |
| Hypertension | 13 (12.9) | 7 (13.5) | 6 (12.2) | 1.000 |
| Ischemic heart disease | 3 (3.0) | 1 (1.9) | 2 (4.1) | 0.610 |
| Neurological disorder | 3 (3.0) | 1 (1.9) | 2 (4.1) | 0.610 |
| Psychiatric disorder | 1 (1.0) | 1 (1.9) | 0 (0.0) | 1.000 |
| Other | 15 (14.9) | 8 (15.4) | 7 (14.3) | 1.000 |
| Location of envenomation |  |  |  |  |
| Hand | 77 (76.2) | 36 (69.2) | 41 (83.7) | 0.105 |
| Leg | 41 (40.6) | 26 (50.0) | 15 (30.6) | 0.068 |
| Other | 1 (1.0) | 1 (1.9) | 0 (0.0) | 1.000 |
| Local symptoms |  |  |  |  |
| Pain | 67 (66.3) | 39 (75.0) | 28 (57.1) | 0.063 |
| Swelling | 100 (99.0) | 52 (100.0) | 48 (98.0) | 1.000 |
| # of large joints involved ± CI | $2 \pm 0$ | $2 \pm 0$ | $2 \pm 0$ | 0.324 |
| Any generalized symptom | 57 (56.4) | 51 (98.1) | 6 (12.2) | 0.000 |
| Reduced consciousness | 1 (1.0) | 1 (1.9) | 0 (0.0) | 1.000 |
| Sweating | 10 (9.9) | 10 (19.2) | 0 (0.0) | 0.001 |
| Hypotension | 19 (18.8) | 19 (36.5) | 0 (0.0) | 0.000 |
| Arrhythmia | 8 (7.9) | 8 (15.4) | 0 (0.0) | 0.006 |
| Shortness of breath | 7 (6.9) | 6 (11.5) | 1 (2.0) | 0.113 |
| Desaturation | 3 (3.0) | 3 (5.8) | 0 (0.0) | 0.243 |
| Abdominal pain | 23 (22.8) | 23 (44.2) | 0 (0.0) | 0.000 |
| Nausea | 33 (32.7) | 28 (53.8) | 5 (10.2) | 0.000 |
| Vomiting | 26 (25.7) | 25 (48.1) | 1 (2.0) | 0.000 |
| Laboratory values on admission |  |  |  |  |
| WBC ( $\times 10^3/\mu\text{l}$) n=98 | $8.5 \pm 0.2$ | $9.2 \pm 0.2$ | $7.7 \pm 0.2$ | 0.008 |
| Platelets ( $\times 10^3/\mu\text{l}$) n=98 | $236 \pm 96$ | $246 \pm 165$ | $225 \pm 147$ | 0.092 |
| Creatinine (mg/dL) n=96 | $0.97 \pm 0.00$ | $1.00 \pm 0.00$ | $0.95 \pm 0.00$ | 0.056 |

|  |  |  |  |  |  |
| --- | --- | --- | --- | --- | --- |
| INR | n=97 | 1.14±0.01 | 1.07±0.00 | 1.21±0.02 | 0.672 |
| Fibrinogen (mg/dL) | n=35 | 259±177 | 257±473 | 263±551 | 0.268 |
| Admission times (minutes) - median [IQR] |  |  |  |  |  |
| Envenomation to hospital |  | 45 [30-61] | 47 [30-78] | 43 [33-60] | 0.703 |
| Envenomation to ICU admission |  | 183 [127-260] | 169 [108-278] | 183 [135-209] | 0.939 |
| Hospital to ICU admission |  | 121 [81-179] | 133 [75-197] | 110 [90-169] | 0.793 |
| Envenomation to antivenin |  |  | 119 [60-237] |  |  |
| Hospital admission to antivenin |  |  | 48 [24-222] |  |  |
| Envenomation to last antivenin |  |  | 195 [110-324] |  |  |

Envenomation primarily occurred between May and November, with a small number of envenomations in the off-season and a peak at the beginning and towards the end of the season. It should be noted that the peak towards the end of the season primarily involved envenomations not requiring antivenin treatment (Fig 2).

**Fig 2.**
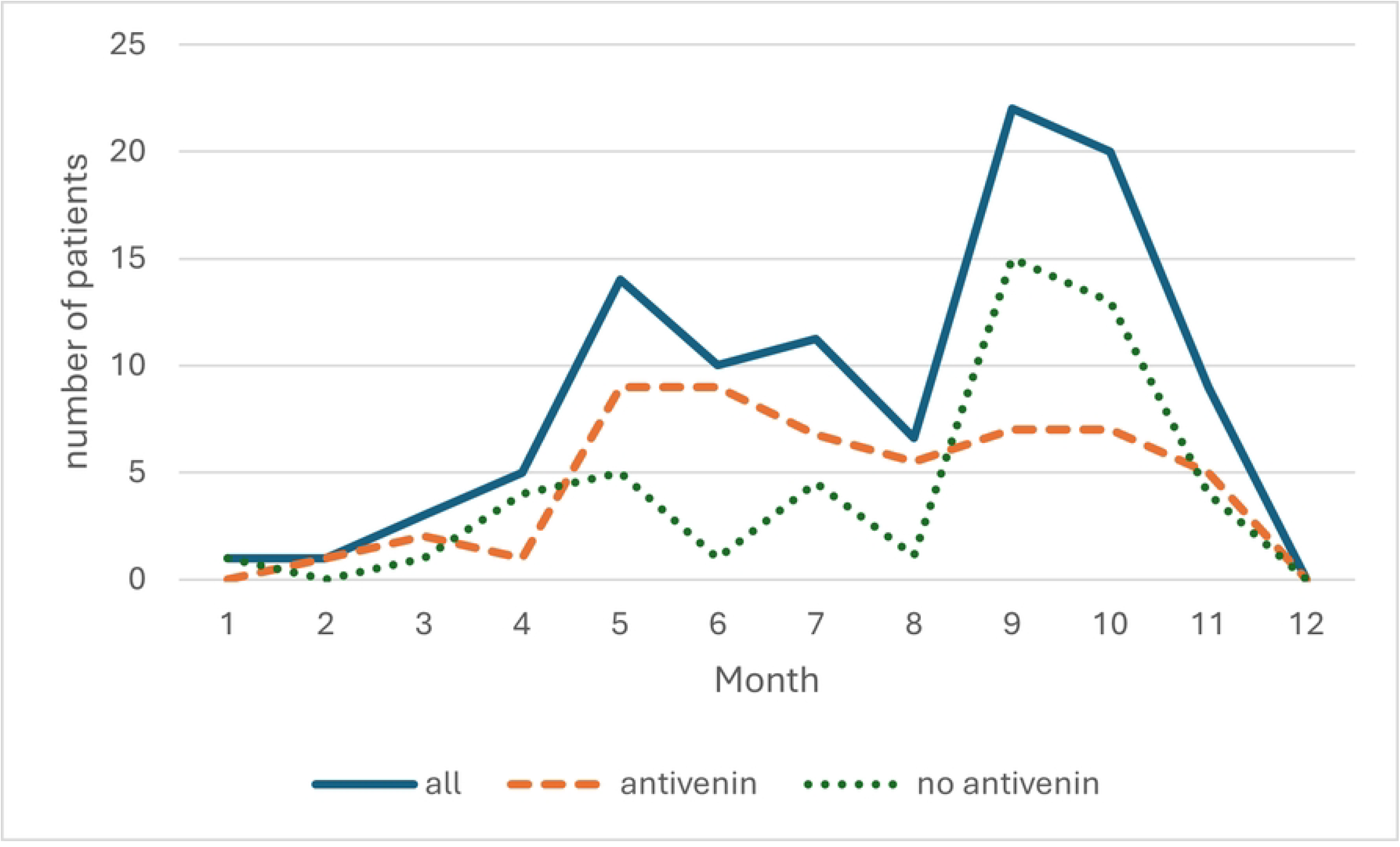
Seasonal envenomation variation. Note: due to missing data from July-August 2023 this data has been interpolated.

Envenomation occurred in the hand in 77 of 101 patients (76.2%), in the leg in 41 patients (40.6%) with 1 patient receiving a bite on the scalp. 18 patients received snake bites in multiple locations. 67 of 101 patients (66.3%) experienced pain at the location of the bite and 100 of 101 (99.0%) patients experienced local swelling. No differences were noted in bite location or local symptoms between those who received antivenin and those who did not (Table 1).

57 of 101 patients (56.4%) experienced one or more generalized symptoms. 51 of these received antivenin as per institutional protocol. 7 protocol deviations occurred, in accordance with clinical judgement of the treating physician, with 1 patient without generalized symptoms receiving antivenin and 6 patients with mild generalized symptoms not receiving antivenin (Table 1). After chart review, the latter deviations were due to clinical judgement attributing symptoms to treatment, rather than to envenomation.

White blood cell count (WBC) on admission of the antivenin group was increased versus the non-antivenin group – 9.2 ± 0.2 x 10^3^/μl versus 7.7 ± 0.2 x 10^3^/μl (p=0.008). Platelet and creatinine levels on admission were also increased in the antivenin versus the non-antivenin group – 246 ± 165 x 10^3^/μl versus 225 ± 147 x 10^3^/μl platelets and 1.00 ± 0.00 and 0.95 ± 0.00 creatinine. However, this did not reach statistical significance (p=0.092 and p=0.056, respectively). Mean international normalized ratio (INR) and mean fibrinogen for the entire population on admission were 1.14±0.01 and 259±177, respectively, with no significant differences noted between groups.

Median time between envenomation and hospital admission was 45 [30-61] minutes, with 121 [81-179] minutes from hospital to ICU admission and 183 [127-260] minutes between envenomation and ICU admission. No differences in admission times were noted between groups. In the antivenin group, median time between envenomation and initial antivenin treatment was 119 [60-237] minutes, with a median of 48 [24-222] minutes elapsing between hospital admission and antivenin treatment. Median time from envenomation to last antivenin treatment was 195 [110-324] minutes (Table 1). Some patients developed symptoms late, thus the maximum time between envenomation and antivenin administration was 1073 minutes. Other patients developed recurrent symptoms despite early antivenin administration; thus, the maximum time between envenomation and last antivenin administration was 3860 minutes.

During the course of their hospitalization, patients who were treated with antivenin received a median of 15 [10-22.5] ml of antivenin in a median of 1 [1-2] administrations (Table 2).

**Table 2.** Treatment and course of hospitalization. ICU: intensive care unit; INR: international normalized ratio; IQR: interquartile range; WBC: white blood cells.

| Variable | All patients<br>n=101 | Antivenin n=52 | No antivenin<br>n=49 | p-value |
| --- | --- | --- | --- | --- |
| Total antivenin dose (ml) - median [IQR] |  | 15 [10-22.5] |  |  |
| Total antivenin treatments - median [IQR] |  | 1 [1-2] |  |  |
| Other treatments received - n (%) |  |  |  |  |
| Vasoactive medication | 7 (6.9) | 7 (13.5) | 0 (0.0) | 0.013 |
| Supplemental oxygen | 5 (5.0) | 5 (9.6) | 0 (0.0) | 0.057 |
| Invasive ventilation | 2 (2.0) | 2 (3.8) | 0 (0.0) | 0.495 |
| Blood products | 2 (2.0) | 2 (3.8) | 0 (0.0) | 0.495 |
| Antiemetics | 19 (18.8) | 15 (28.8) | 4 (8.2) | 0.010 |
| Non-opiate analgesia | 50 (49.5) | 35 (67.3) | 15 (30.6) | 0.001 |
| Opiate analgesia | 41 (40.6) | 28 (53.8) | 13 (26.5) | 0.008 |
| Surgical intervention | 4 (4.0) | 3 (5.8) | 1 (2.0) | 0.618 |
| Laboratory values - median [IQR] |  |  |  |  |
| WBC max ( x 10<sup>3</sup>/μl) | 9.6 [7.9-11.9] | 11.1 [9.2-13.4] | 8.4 [7-10.1] | 0.001 |
| Platelets nadir ( x 10<sup>3</sup>/μl) | 175 [133-213] | 151 [118-188] | 186 [161-240] | 0.002 |
| Creatinine max (mg/dl) | 0.96 [0.85-1.1] | 0.99 [0.86-1.12] | 0.95 [0.85-1.05] | 0.091 |
| INR max | 1.06 [1.02-1.15] | 1.09 [1.04-1.18] | 1.03 [1.01-1.11] | 0.566 |
| Fibrinogen nadir (mg/dl) | 241 [206-289] | 235 [201-293] | 252 [231-285] | 0.414 |
| Outcomes - median [IQR] |  |  |  |  |
| ICU LOS | 2 [2-3] | 3 [2-3] | 2 [1-2] | 0.001 |
| Hospital LOS | 3 [2-4] | 4 [3-5] | 3 [2-4] | 0.001 |

26/52 (50.0%) patients receiving antivenin required only 10 ml antivenin (one dose according to institutional protocol), with 13/52 (25.0%) requiring 20 ml and 13/52 (25.0%) requiring more than 20 ml. The maximum dose administered was 70 ml, which was received by 1 patient in 3 separate administrations. 2 patients developed an early immune reaction to antivenin, with one of these developing anaphylaxis requiring invasive ventilation.

Besides antivenin, 7/101 patients (6.9%) required vasoactive medication, 5 (5.0%) required supplemental oxygen and, of these, 2 (2.0%) required invasive ventilation. 2 (2.0%) required blood products. All these were in the group that received antivenin. Additionally, 15/52 (28.8%) in the antivenin group and 4/49 (8.2%) in the non-antivenin group, were treated with antiemetics (p=0.010), 35/52 (67.3%) and 15/49 (30.6%) – respectively – required non-opiate analgesia (p<0.001), 28/52 (53.8%) and 13/49 (26.5%) required opiate analgesia (p=0.008) and 3/52 (5.8%) and 1/49 (2.0%) received surgical intervention (p=0.618); surgical intervention was carried out in deviation from standard protocol [1, 12].

In terms of laboratory values, mildly increased maximal leukocytes were noted in the antivenin group relative to the non-antivenin group (median 11.1 [9.2-13.4] x 10^3^ per μl versus 8.4 [7.0-10.1] x 10^3^ per μl, p<0.001). A reduction in platelets was noted in most patients over the course of the hospitalization, with 49/52 (94.2%) patients in the antivenin group and 33/49 (67.3%) of patients in the non-antivenin group developing a platelet reduction relative to admission (p=0.001), with a median nadir of 151 [118-188] x 10^3^ per μl in the antivenin versus 186 [161-240] x 10^3^ per μl in the non-antivenin group, p<0.001. 3 patients in the antivenin group developed a severe thrombocytopenia of less than 50 x 10^3^ per μl, with no cases of severe thrombocytopenia in the non-antivenin group. No patients developed major bleeding or required platelet transfusion.

No substantial pathologies or differences in coagulation factors were noted between groups, with a median maximal INR of 1.06 [1.02-1.15] and a median fibrinogen nadir of 241 [206-289] mg/dl for the cohort. One patient presented with an INR of 10.4, which was thought to be due to laboratory error and, in fact, normalized without treatment upon repeat collection. One patient on chronic warfarin anticoagulation presented with an INR of 3.49 and another patient developed an INR of 1.88 during course of hospitalization which normalized without treatment. None of these patient received antivenin or blood products. No patient developed clinically significant fibrinogen abnormalities.

Median maximal creatinine was 0.96 [0.85-1.1] with no differences between groups. No patient developed acute kidney injury during their hospitalization.

Median length of stay in the ICU was 3 [2-3] days for the antivenin and 2 [1-2] days for the non-antivenin group (p<0.001). Median hospital length of stay was 4 [3-5] days for the antivenin group and 3 [2-4] days for the non-antivenin group (Table 2). No patients died during the course of their hospitalization.

## Discussion

The primary and most effective treatment of snake bite envenomation, including that resulting from *D. palaestinae* bite, continues to be administration of specific or polyvalent snake antivenin, which drastically reduces mortality and morbidity [8, 9]. However, antivenin administration is costly, with a single fixed dose regimen estimated cost of $7000, with repeat administration sometimes required [4, 5, 11]. Furthermore, certain antivenins have historically been in short supply [11]. These limitations can form substantial barriers to treatment access, especially in limited-resource settings. Moreover, antivenin administration can itself be associated with substantial morbidity, including delayed reactions up to a month after administration [4, 8, 10]. Thus, it is important to define the minimal necessary dose for effective reversal of envenomation, as well as to define patient populations who can be safely treated without antivenin administration.

This study of 101 patients diagnosed with *D. palaestinae* envenomation is the largest published cohort of human *D. palaestinae* envenomations to date. It was designed to evaluate an envenomation treatment protocol that, in contrast with the standard fixed-dose regimen, was developed in order to minimize the use of antivenin, thus minimizing side effects and overall morbidity, reducing costs and reducing antivenin use, especially in periods of scarcity [10]. In this protocol, local symptoms were treated conservatively, without use of antivenin [5, 6, 8] and treatment of both mild and severe systemic symptoms employed dose scaling antivenin, with an initial 10 ml dose of antivenin and repeat dosing in patients with ongoing systemic symptoms, in accordance with treatment response. In our study, 52 of 101 (51.5%) patients received antivenin, with 26 of those receiving 10 ml of antivenin and another 13 receiving 20 ml. This is in contrast with the standard protocol, in which most or all of these patients would have received 50 ml of antivenin [4, 5, 11]. This protocol resulted in an estimated cost saving of over $500,000, as well as an estimated 2 fewer allergic reactions, including 1 less anaphylaxis, with no increased morbidity or mortality related to the reduced dose protocol.

In the current study, nearly all patients diagnosed with *D. palaestinae* envenomation develop swelling and a majority suffering from site pain, with a higher percentage of pain reported among those requiring antivenin. We also report abdominal symptoms, respiratory distress, hypotension and with 1 case of reduced consciousness associated with envenomation. This is similar to previous studies [4, 8-10]. Unlike those studies, our patients did not develop convulsions, coagulopathy or necrosis, possibly due to early antivenin treatment when necessary, with a median time between envenomation and initial antivenin treatment of 119 [60-237] minutes. Similar to previous studies, a majority of our cohort showed a reduction in platelets over the course of the hospitalization [13]. This was true whether or not the patient was treated with antivenin (though patients who received antivenin had a more substantial platelet reduction), and, similar to some [14], but unlike other [15], reports in other Viperidae species, there was no apparent response to antivenin. This may be due to venom-induced macrophage and hepatocyte sequesterization and clearance of platelets, as shown in other species of Viperidae [16, 17].

Similar to previous studies [9], we show that some patients show a latent development of symptoms and repeat symptom occurrence. Thus the maximum time between envenomation and first antivenin administration was 17.8 hours and the maximum time until last antivenin administration (due to repeat symptoms) was 64.3 hours.

This study has several limitations. Firstly, it is a retrospective data analysis of an unmatched cohort, and, therefore, its conclusions should be approached with caution. Furthermore, though a comparison has been made between those who did and did not receive antivenin, due to the inherent clinical differences between these groups, no true control group was employed. Rather, in order to evaluate the antivenin protocol of the current study, historical controls from the scientific literature were employed, which also has substantial limitations. Nevertheless, we demonstrated no morbidity or mortality related to the protocol.

## Conclusions

In this cohort of 101 patients with *D. palaestinae* envenomation we show that a dose-scaling antivenin protocol can be safely used in the treatment of envenomation, thus reducing morbidity related to antivenin administration as well as reducing costs. This suggested the need for a randomized control trial comparing a fixed dose regimen to the escalation protocol described in this study, as well as suggest the development of similar sliding scale protocols for envenomations due to other snake species.

## Data Availability

All data files are available from the Open Science Foundation database (accession number osf.io/2p685).

